# Development and Validation of the Michigan Chronic Disease Simulation Model (MICROSIM)

**DOI:** 10.1101/2022.03.21.22271857

**Authors:** James F. Burke, Luciana L. Copeland, Jeremy B. Sussman, Rodney A. Hayward, Alden L. Gross, Emily M. Briceño, Rachael Whitney, Bruno J. Giordani, Mitchell S. V. Elkind, Jennifer J. Manly, Rebecca F. Gottesman, Darrell J. Gaskin, Stephen Sidney, Kristine Yaffe, Ralph L. Sacco, Susan R. Heckbert, Timothy M. Hughes, Andrzej T. Galecki, Deborah A. Levine

**Author notes:** **Corresponding Author:** James F. Burke, MD, MS, The Ohio State University Wexner Medical Center, Department of Neurology, Room 734, 395 w 12^th^ Ave, Columbus OH, 43210.

## Abstract

Strategies to prevent or delay Alzheimer’s disease and Alzheimer’s disease-related dementias (AD/ADRD) are urgently needed. Blood pressure (BP) management is a promising strategy for AD/ADRD prevention and the key element in the primary and secondary prevention of atherosclerotic cardiovascular disease (ASCVD), yet the effects of different population level BP control strategies across the life course on AD/ADRD are not known.

Large-scale randomized controlled trials are the least biased approach to identifying the effect of BP control on AD/ADRD, yet trials may be infeasible due to the need for prolonged follow-up and very large sample sizes. Thus, simulation analyses leveraging the best available observational data may be the best and most practical approach to answering these questions.

In this manuscript, we describe the design principles, implementation details, and population-level validation of a novel population health microsimulation framework, the MIchigan ChROnic Disease SIMulation (MICROSIM), for The Effect of Lower Blood Pressure over the Life Course on Late-life Cognition in Blacks, Hispanics, and Whites (BP COG) study of the effect of BP levels over the life course on cognitive decline and dementia.

MICROSIM was designed by applying computer programming best practices to create a novel simulation model. The initial purpose of this extensible, open-source framework is to explore a series of questions related to the impact of different blood pressure management strategies on late-life cognition and all-cause dementia, as well as the effects on race differences in all-cause dementia incidence. Ultimately, though, the framework is designed to be extensible such that a variety of different clinical conditions could be added to the framework.

## Introduction

Alzheimer’s disease and Alzheimer’s disease-related dementias (AD/ADRD) are major causes of death and disability in older individuals. Preventing or delaying AD/ADRD can lead to better survival, less disability, less nursing home use, lower health care costs, and better quality of life. Blood pressure (BP) management is a promising strategy for AD/ADRD prevention and the key element in the primary and secondary prevention of atherosclerotic cardiovascular disease (ASCVD). Little is known about the effect of BP treatment on the combined outcomes of ASCVD and AD/ADRD at the population level.

Accumulating evidence over the past 20 years has led to important new clinical guidelines for more aggressive treatment of modifiable vascular risk factors. Most policy assessments and simulation models informing these new guidelines mainly, or solely, consider BP’s impact on hearts attacks and strokes, but not AD/ADRD.^1–4^ Yet BP is a strong risk factor for AD/ADRD, and consideration of BP’s effect on AD/ADRD and other disease states may influence “optimal” treatment. For example, while lowering BP to optimal levels (<120/80 mm Hg) reduces CVD events, mild cognitive impairment (MCI), and the combination of MCI and AD/ADRD in individuals with high CVD risk,^5,6^ this group in the US is relatively small. It is unclear whether lowering BP to optimal levels also reduces CVD and AD/ADRD in the larger group of adults at lower CVD risk (e.g., Black individuals age 55 with systolic BP 130-139 mmHg). If higher BP treatment intensity early in /the life course has a large effect on late-life cognition and AD/ADRD, then the optimal timing, treatment threshold, and intensity of BP treatment initiation should shift to earlier ages and more intense treatment. Similarly, estimating the independent effects of BP treatment intensity on CVD vs. AD/ADRD may enable more accurate characterization of quality of life, cost-effectiveness and societal benefit and thus inform optimal policy.

While large-scale randomized controlled trials are the least biased approach to addressing these questions, such trials may be infeasible. Thus, simulation analyses leveraging the best available observational data may be the best and most practical approach to answering these questions. In this manuscript, we describe the design principles, implementation details, and population-level validation of a novel population health microsimulation framework, the MIchigan ChROnic Disease SIMulation (MICROSIM), for The Effect of Lower Blood Pressure over the Life Course on Late-life Cognition in Blacks, Hispanics, and Whites (BP COG) study of the effect of BP levels over the life course on cognitive decline and dementia. MICROSIM was designed by applying computer programming best practices to create a novel simulation model. The initial purpose of this extensible, open-source framework is to explore a series of questions related to the impact of different blood pressure management strategies on late-life cognition and all-cause dementia, as well as the effects on race differences in all-cause dementia incidence. Ultimately, though, the framework is designed to be extensible such that a variety of different clinical conditions could be added to the framework.

## Methods

### Rationale for Using a Simulation Approach

Randomized controlled trials (RCTs) are the optimal study design to determine the efficacy of different BP treatment algorithms (e.g., treatment intensity, the timing of initiation) and patient selection strategies on combined cognitive and vascular outcomes. However, it is challenging to design RCTs for these questions. Ideally, we would want evidence from large, long-term RCTs to address the effect of mid-life blood pressure treatment on late-life cognition, but such trials would be at best cumbersome and costly and, at worst infeasible. Such trials would require enormous sample sizes, rigorous case capture, and robust intervention fidelity — all maintained across decades. Further, even if such a trial is eventually conducted, meaningful results would likely be decades off — during which time the treatment paradigm may have evolved such that the results are no longer meaningful. Even if these problems could be addressed, these studies would not be able to inform patient care in the near term. Shorter-term trials are more feasible but cannot directly determine how short-term treatment decisions influence long-term outcomes since one would have to make assumptions about the importance of short-term surrogate measures and/or make extrapolations from higher to lower risk populations.

Simulation analyses may be able to provide guidance when clinical trials are infeasible and interim guidance for important clinical questions while waiting for the results of better, long-term trials. Simulations can combine the strengths of the best available data sources. First, by using the best available longitudinal observational data they can account for complex natural histories and competing risks. Second, by incorporating the best inferences regarding treatment effectiveness (trial-based when available, and the best observational estimates^7^ when trials are not available) they can credibly account for a range of treatments effects. By leveraging these strengths in a carefully-specified framework, simulations may be the best tools to estimate long-term treatment effects, particularly in low-risk populations. Simulations can also explore the societal consequences of clinical and policy interventions and inform the likely relative yields of such policies.

Yet, simulations suffer multiple limitations. Simulations’ intrinsic complexity often makes them seem like “black box science,” frequently leading to a lack of transparency and concerns about the accuracy of their results. Moreover, most current models are built individually and fail to build upon one another, limiting progress and potentially amplifying concerns regarding accuracy. Creating the framework for an effective simulation can be complicated, time-intensive, and require the careful evaluation of many individual decisions. Here, we describe our approach to the development of a simulation model and our attempt to mitigate these limitations.

### Simulation Design Principles

Simulation analyses are implemented by developing software codebases that encode the simulation logic and stochastic elements for data creation. These codebases are often large, containing tens of thousands of lines of computer code or more. As codebases increase in size, so does the expected number of bugs.^8^ Ensuring that simulation analyses produce accurate and reliable results requires sound software development processes such that the code implements the underlying simulation logic with sufficient code quality. To this end, we developed a series of principles and strategies based on computer programming best practices to guide MICROSIM development: Transparency, Readability, Simplicity, Testing, and Validation^9^.

1. Transparency — the mere possibility that other groups will assess one’s codebase incentivizes coders to be particularly careful and make sure the code is readable and coherent. The entire MICROSIM codebase^10^, including the notebooks used to develop MICROSIM’s inputs and the notebooks used for the validation analyses^11^ presented in this manuscript, are all publicly available on GitHub^10^. Opening the repository to other investigators also broadens the user base and thereby increases the likelihood of identifying errors in the simulation, if present.
2. Readability — the more readable the code, the less likely it is for errors to emerge. We used three main strategies to achieve this. First, we hired a software developer with industry experience (LC, Ann Arbor, MI) to assist with the coding and the software development methodology. Second, the core elements of the simulation were developed via pair programming, where two coders (JB and LC) sat side-by-side and shared a keyboard to develop key simulation elements. Third, we used automatic formatting tools (Black^12^) that ran when a file was saved to have a consistent and predictable code style, making the code easier to read and write.
3. Simplicity — the simplest possible model structures and assumptions were employed whenever possible. The simpler the logic to be implemented, the less likely it would be implemented with error. This principle was intended to apply both to the simulation code and overall simulation structure. At the code-level, this principle is largely a statement of priorities. If a relatively simple data structure is able to represent the data, then it should be preferred over a more complex data structure, even if the complexity may, for example, improve performance. Similarly, while more complex statistical models may represent the data structure modestly better, unless there are major gains in performance, simpler statistical models should be preferred as they are less likely to be implemented with error.
4. Testing — in a large codebase, it is possible that a change in one area may have unintended consequences elsewhere. We employed unit tests in a largely test-driven development paradigm to mitigate this risk.^13^ That is, for a new piece of code or bug fix, first, a test was developed that would fail until the code was correctly implemented, and that same test would only pass when the code was implemented correctly. Tests also ran automatically on TravisCI^14^ when new code was pushed to GitHub, making it easy to see if a change caused a test to fail.
5. Validation against the best available data— the core element to ensure accurate simulation results was to compare the simulation results to real world data that were not included in the model derivation whenever possible. This manuscript summarizes the key validation steps that MICROSIM has undergone to date.

### Overview of Simulation Evidence Evaluation Hierarchy and Structure

MICROSIM is an agent-based Monte Carlo simulation using varying regression-based models (e.g. logistic regression for binary outcomes, Cox regression for time-to-event data) to model annual transition probabilities in risk factors and outcomes.^10^ Model assumptions and inputs were derived from the best available evidence. Evidence was selected by applying an evidence hierarchy that prioritized meta-analyses of controlled trials over individual controlled trials over high-quality observational evidence. When none of these were available, we derived regression models from a combination of six large scale cohort studies: Atherosclerosis Risk in Communities Study (ARIC)^15^, Coronary Artery Risk Development in Young Adults Study (CARDIA)^16^, Cardiovascular Health Study (CHS)^17^, Framingham Offspring Study (FOS)^18^, Multi-Ethnic Study of Atherosclerosis (MESA), and Northern Manhattan Study (NOMAS).^19^ MICROSIM is designed to model scenarios for preventing ASCVD and dementia, based on prior simulations to model related questions in ASCVD^2^ and dementia prevention.^20^ **(Figure 1)**

**Figure 1:**
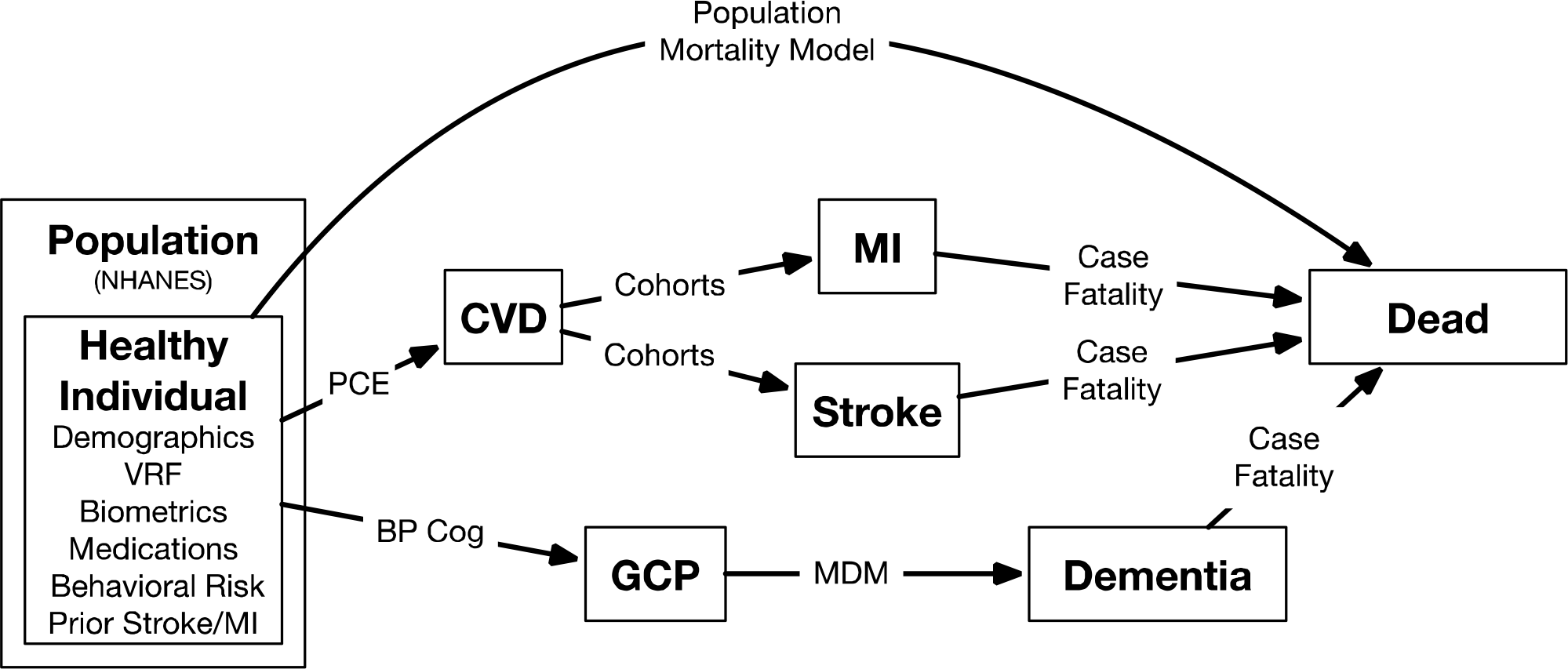
Overview of MICROSIM. NHANES = National Health and Nutrition Examination Survey VRF = Vascular Risk Factors PCE = Pooled Cohort Equations BP Cog = Study to relate Blood Pressure to cognition, “The Effect of Lower Blood Pressure over the Life Course on Late-life Cognition in Blacks, Hispanics, and Whites (BP-COG)” GCP = Global Cognitive Performance MDM = Michigan Dementia Model

In MICROSIM, we examine how healthy individuals transition into non-ASCVD death, fatal or non-fatal ASCVD, all-cause dementia, or remain free of all those events. MICROSIM updates the status of healthy individuals annually using Monte Carlo methods. The MICROSIM population is derived from the nationally representative National Health and Nutrition Examination Survey (NHANES). Risk factors, cognition, ASCVD events, non-ASCVD death, and all-cause dementia transition rates are primarily estimated using predictive models derived from large pooled cohort studies. Treatment effects are derived from randomized trials as described in the “Simulation Details” sections below. Based on existing research, quality-adjusted life years (QALYs) are assigned to each state.

### Simulation Details— Baseline Population^21^

Our goal was to create a population representing the overall United States (US) population while also focusing on specific (e.g., condition-specific) subgroups. We built our baseline population using adult (18+) data from NHANES. NHANES is a nationally representative serial cross-sectional survey designed to assess the health and nutritional status of the US population and has been continuously assessing demographics, vascular risk factors, and vascular events since 1999 using consistent methodology.^22^ NHANES does not repeatedly measure cognitive data and thus cognitive model inputs were drawn from different data sources. Each simulation run is initiated with data representing a specific national sample based on a given NHANES wave and then advanced forward over time.

We built a dataset of relevant NHANES risk factors for CVD or all-cause dementia from 1999-2017, including demographics (age, sex, race/ethnicity, education (less than high school, some high school, high school graduate, some college, college graduate), vascular risk factors (systolic and diastolic BP [SBP, DBP], total cholesterol, triglycerides, low-density lipoprotein [LDL] cholesterol, high-density lipoprotein [HDL] cholesterol, glycosylated hemoglobin [HgbA1C]), biometrics (body mass index [BMI], waist circumference), behaviors relevant to vascular risk (physical activity, smoking), medications (anti-hypertensive agents, statins, other lipid-lowering agents) and vascular events (stroke, myocardial infarction (MI))^21^. We then used multiple imputations with chained equations to account for missing data. Imputation strategies for a given variable included all other variables as covariates with the following exceptions: triglycerides and LDL, which were excluded from models for each other due to collinearity; and anti-hypertensive medications. Imputation models for anti-hypertensive medications added the following: an indicator variable for whether the patients’ BP qualified as hypertensive by the Eighth Joint National Committee (JNC-8) criteria, interactions between DBP, SBP, and JNC-8 hypertension, and interactions between SBP, DBP, and self-reported hypertension to improve predictiveness. The simulation is initialized by defining a starting year (1999-2017) and population size. These parameters are used to select the baseline MICROSIM population from the fully imputed dataset using survey-weighted selection with replacement.

### Simulation Details —Change in Risk Factors over Time^23^

We built longitudinal regression models to predict each risk factor in the baseline population as the dependent variable using pooled individual participant data from the six cohorts (ARIC, CARDIA, CHS, FOS, NOMAS). Predictor variables included race/ethnicity, smoking, gender, lagged (i.e., the value of the factor at the immediately prior time point), and mean lagged (i.e., mean value of the factor across all prior time points) fixed effects for the vascular risk factor of main interest and for all other time-varying risk factors. Regression results were stored as JavaScript Object Notation (JSON) files, including characteristics of the residual distributions (mean and standard deviation). Advancing in one-year increments, new values of each risk factor were calculated for each individual by summing the linear predictor from the corresponding regression model with a random draw from the residual distributions. SBP and DBP were log-transformed throughout to improve model fit and de-transformed when estimating updated individual factors.

### Simulation Details — Cardiovascular Events

For each time step in the model, each individual’s risk of a CVD event was calculated based on their vascular risk factors using the updated Pooled Cohort Equation (PCE), which predicts the risk of stroke, MI or vascular death.^24^ To assign events, a random uniform number on the interval of [0, 1) was generated for each person, for each year. Individuals, then, were each assigned an event if their random uniform number was less than their estimated risk and were not assigned an event otherwise.

After determining whether or not individuals would have a CVD event, specific event types were assigned using an event-partitioning model^25^. Using individuals with a stroke or MI in the combined cohorts, a logistic regression model was built that included factors understood to differentially predict stroke vs. MI: age, lagged SBP, DBP, BMI, triglycerides, HgbA1C, gender, and race/ethnicity. In the simulation, when an individual was assigned a cardiovascular event, the inverse logit of the linear predictor of the partitioning model was used to determine the probability that the event would represent a stroke. Event types were randomly assigned while maintaining the stroke vs. MI risk distribution by determining whether a [0, 1) random uniform number was below that partition threshold, assigning a stroke if so or an MI if not. Fatal vs. non-fatal CVD event determinations were made stochastically by determining whether another [0, 1) random uniform number was less than the case fatality estimates for stroke (0.15)^26–29^ and MI (0.13).^30^

### Simulation Details — Non-cardiovascular (non-CV) Mortality^31^

After assigning cardiovascular events, non-CV mortality was assigned using a similar approach. The risk of non-CV mortality was estimated from a Cox proportional hazards model developed in NHANES. Specifically, long-term mortality data was linked to the combined NHANES dataset, and underlying causes of death were identified. A Cox-proportional hazards model was developed in this dataset predicting time to non-CV death after adjusting for age, age squared, race/ethnicity, gender, mean SBP, mean DBP, HgbA1C, total cholesterol, triglycerides, BMI, and smoking status. To estimate individual non-CV mortality risk, the baseline cumulative hazard function was approximated with a quadratic function. The sum of the products of baseline covariates and regression coefficients was used to estimate the individual-level linear predictor (xb) of non-CV mortality for each individual. The cumulative hazard of death was estimated at time t and t-1 by taking the product of the cumulative hazard at each time and the exponentiated linear predictor (cumulative hazard * e^xb^) and the risk of mortality in a given year was taken as the difference in cumulative hazard between the two time points. For each individual, in each year, a random uniform number on the interval of 0 to 1 was drawn, and if this was less than the estimated risk of non-CV death, the patient was assigned non-CV death; otherwise the patient continued in the simulation alive into the next wave.

### Simulation Details — Cognition and Dementia

In prior work, our group built longitudinal models to predict global cognitive performance (GCP) using pooled data from the pooled cohorts containing cognitive measures.^32,33^ In brief, trained cohort staff administered cognitive function tests longitudinally in-person to participants using cognitive tests. To make inferences about cognitive domains instead of individual cognitive test items, and to resolve the challenge of different cognitive tests administered across the cohorts, we co-calibrated available cognitive test items into a factor representing global cognition (global cognitive performance), using item response theory methods (a graded response model) that leverage all available cognitive information in common across cohorts and test items unique to particular cohorts.^34–37^ GCP factor scores were estimated using the regression-based method in Mplus, such that a 1-point difference represents a 0.1 SD difference in the distribution of cognition across the cohorts. Higher cognitive scores indicate better performance.

Each individual in MICROSIM was assigned a baseline GCP random effect by sampling with replacement from the distribution of GCP random effects in the published model. We then estimated each individual’s future GCP values based on the individual’s linear prediction from the model, the individual’s random effect, and a random draw from the overall residual distribution.

To stratify dementia risk across risk factors, we built a Cox proportional hazards model in ARIC, CHS and FOS to predict all-cause dementia, including baseline values of GCP, education, age, sex, race/ethnicity, as well as GCP slope (change in GCP scores over time) as covariates. ^38^, CHS^39^, and ^40^measured incident dementia by physician-adjudication using standard diagnostic criteria, study-specific protocols, and all available data including in-person neuropsychological and neurologic assessments, telephone interviews (participant or informant), brain imaging, and medical record review. Covariates are summarized in **Table 1**.

**Table 1:** Coefficients for Cox Model predicting time to all-cause dementia using combined cardiovascular cohort data.

|  | **HR [95% CI]** |
| --- | --- |
| Baseline GCP (per one unit of Baseline GCP) | 0.927 [0.92-0.935] |
| GCP Slope (per one unit change in GCP per year) | 0.999 [0.999-0.999] |
| Baseline age (years) | 1.108 [1.101-1.114] |
| Female | 1.1 [0.995-1.215] |
| Education (ref: College Graduate or Higher) |  |
| Eighth Grade or Less | 1.032 [0.85-1.252] |
| Some High School | 1.088 [0.919-1.287] |
| Completed High School/GED | 0.919 [0.807-1.045] |
| Some College but no degree | 0.797 [0.678-0.939] |
| Non-Hispanic Black (ref: Non-Hispanic White) | 1.214 [1.048-1.406] |

GCP = Global Cognitive Performance

The incidence of dementia is somewhat higher in the combined cohorts than has been observed in a prior meta-analysis of epidemiologic studies of dementia incidence and summarized in an equation by Brookmeyer et al.,^41,42^ likely due to slightly different definitions of dementia. Thus, we fit a quadratic function to the baseline survival curve from the cohort-derived Cox model and searched parameter space for modifications of the quadratic parameters such that the final dementia incidence most closely fit the Brookmeyer equation. This recalibration can easily be “turned off” for a given analysis if the combined cohort incidence of dementia was thought to represent dementia incidence more accurately in the target population.

### Simulation Details — Treatment Effects

A central goal of this simulation is to estimate how CVD events are impacted in the counterfactual where BP treatment is more or less intense than usual care. This goal requires reliable estimates of how an additional anti-hypertensive medication impacts risk.

MICROSIM uses clinical trial effect size estimates whenever available but for logistical reasons, does so by recalibrating initial estimates from observational studies. In observational analyses, the association between BP levels and CVD events generally under-estimates the CVD treatment effect (i.e., relative risk [RR] reduction) observed for anti-hypertensive medications in trials.^43^ That is, an anti-hypertensive medication in a trial may lower BP by 5/3 mm Hg and have a RRR for ASCVD of 0.2. However, in observational data, individuals with BPs that are 5/3 mm Hg lower will have a smaller RRR for ASCVD, 0.1. Therefore, we modeled anti-hypertensive treatment effects by first applying the mean BP lowering observed across anti-hypertensive trials for each anti-hypertensive agent added — 5.5/3.1 mm Hg.^44^ To make ASCVD treatment effects consistent with the superior clinical trial evidence, a treatment recalibration phase at the end of each annual increment adjusted for the smaller treatment effect of an additional anti-hypertensive medication on ASCVD observed in clinical trials based on the trial/observational RR reduction per achieved mm Hg BP reduction (RR of 0.79/BP medication for stroke and RR of 0.87/BP medication for MI) by randomly rolling back CVD events. Individuals who received an additional anti-hypertensive medication were randomly chosen to have their events rolled back, weighted by their inverse untreated risk such that the highest risk patients would be least likely to be chosen. ^29^

### Simulation Details — Quality Adjusted Life Years (QALYS)

QALYs were assigned to each individual in each iteration. If an individual had no events (no MI, stroke, or dementia), they were assigned an average QALY based on age.^45^ For events (stroke, MI, all-cause dementia), baseline age-based utilities were reduced by relative effects for each age. For stroke and MI, the magnitude of the reduction varied such that the effect was greater in the year of an incident event (RR, 0.67 for stroke, RR, 0.88 for MI) and lower in subsequent years (RRs, 0.9 for both stroke and MI).^2^ For dementia, we used a multiplier of 0.80 for the first year of incident dementia with a 0.01 reduction in each subsequent year. (i.e. 5 years after a dementia, multiplier = 0.75).^46^

## Codebase Overview

The simulation is structured around two core classes — Person and Population. The Person class largely serves as a data store of risk factors and outcomes with properties and methods for getting and updating the Person’s state throughout the simulation. The Population class manages a group of Persons. Specific Population instances are responsible for loading data from a given data source, advancing the population forward in time (i.e., by one year), recalibrating population-level outcomes, and simple reporting. To help with advancing, the Population uses Models, which compute the next property (e.g., risk factor or outcome) for a Person, which it obtains from a Model Repository, a Model store that can also determine which Model to apply to a Person. Most Models are Regression Models, which take a series of regression coefficients and estimate individual-level risks for a given Person.

Each of these elements is designed to be adaptable and extensible to new populations, new parameters, and new risk models. For example, one could initialize the simulation with novel data, and the Population class could then be subclassed and that logic added to initialized Person objects. Similarly, it is readily possible to change which Models are used to implement the changes in specific factors over time. For example, replacing the existing ASCVD risk model with a newer or different population risk model can be easily accommodated.

## Validation Methods and Results

### Validation Strategy

We sought to identify the key elements that would be most likely to influence the overall model accuracy regarding our core constructs and central research questions. For these elements, then, we identified the best validation strategy. The core elements we focused on were overall population representativeness, CV risk factors over time, CV events, and dementia. Validation of Baseline Simulated Population^47^

**Table 2** compares two simulated populations to published NHANES standards. We found that our randomly selected 500,000 person simulated population nearly identically matched published demographics in the 2007-2010 survey-weighted NHANES cohort.^48^ Similarly, we found excellent matching in demographics and vascular risk factors in our 500,000 person simulated population with hypertension (SBP > 140/90 mm Hg or any anti-hypertensive medication) to a published survey-weighted NHANES cohort of individuals with hypertension in 2013.^49^

**Table 2:** Comparison of Simulation Risk Factors to NHANES and the NHANES sub-population with hypertension.

|  | 2007-2009<sup>37</sup> |  | 2013, Hypertension<sup>38</sup> |  |
| --- | --- | --- | --- | --- |
|  | NHANES | Simulation | NHANES | Simulation |
| **Age in** |  |  |  |  |
| years(mean) | 45.9 | 45.9 | 60.0 | 58.4 |
| % Female | 51.7% | 51.7% | 50.0% | 49.7% |
| **Race Ethnicity** |  |  |  |  |
| % White | 68.4% | 68.3% | 71.0% | 69.6% |
| % Black | 11.5% | 11.4% | 14.0% | 14.0% |
| % Hispanic | 13.6% | 13.6% | 10.0% | 10.3% |
| **BMI** |  |  |  |  |
| kg/m<sup>2</sup>(mean) | 28.5 | 28.5 | 31.0 | 30.9 |
| **Hypertension** |  |  |  |  |
| (JNC-8) |  |  | 80.0% | 82.5% |
| SBP (mean) |  |  | 133.4 | 132.1 |
| DBP (mean) |  |  | 71.6 | 72.2 |
| **Medications** |  |  |  |  |
| % Anti-hypertension |  |  | 41.0% | 41.4% |
| % Statin |  |  | 41.0% | 41.4% |

### Validation of Vascular Risk Factor Population Over Time^50^

Although we found the simulation framework closely reproduced longitudinal changes in actual cohorts (results not shown), this finding is somewhat circular since those cohorts were used to inform the simulation’s models. We, therefore, assessed how well the simulation framework reproduces longitudinal changes over time by comparing a longitudinal cohort from the simulation to a pseudo-cohort from NHANES (derived from repeated cross-sectional NHANES) as a more robust assessment of the simulation’s fidelity. Specifically, we built a simulated population of 250,000 adults in 1999 and advanced the simulated population for 18 years until 2017 (the most recent year for which NHANES data was available). We then removed simulated individuals that died prior to 2017 from the population. For our NHANES comparator, we included adults age 36 (baseline age 18 + 18 years of age advancement) or older and excluded adults that immigrated into the US as the simulation does not account for in or out migration. A histogram of cardiovascular risk factors in the simulation (initialized in 1999 and advanced 18 years) and the pseudo-cohort (NHANES 2017 without in-migration) is presented in **Figure 2**. The simulation generally closely reproduced both central tendencies and variances of risk factors, except for over-predicting DBP (mean 78.7, SD 9.2 vs. 71.6 mm Hg, SD 10.9) and slightly under-predicting variance in total cholesterol (mean 200.0, SD 31.0 vs. 196.8 mg/dL, SD 41.3).

**Figure 2:**
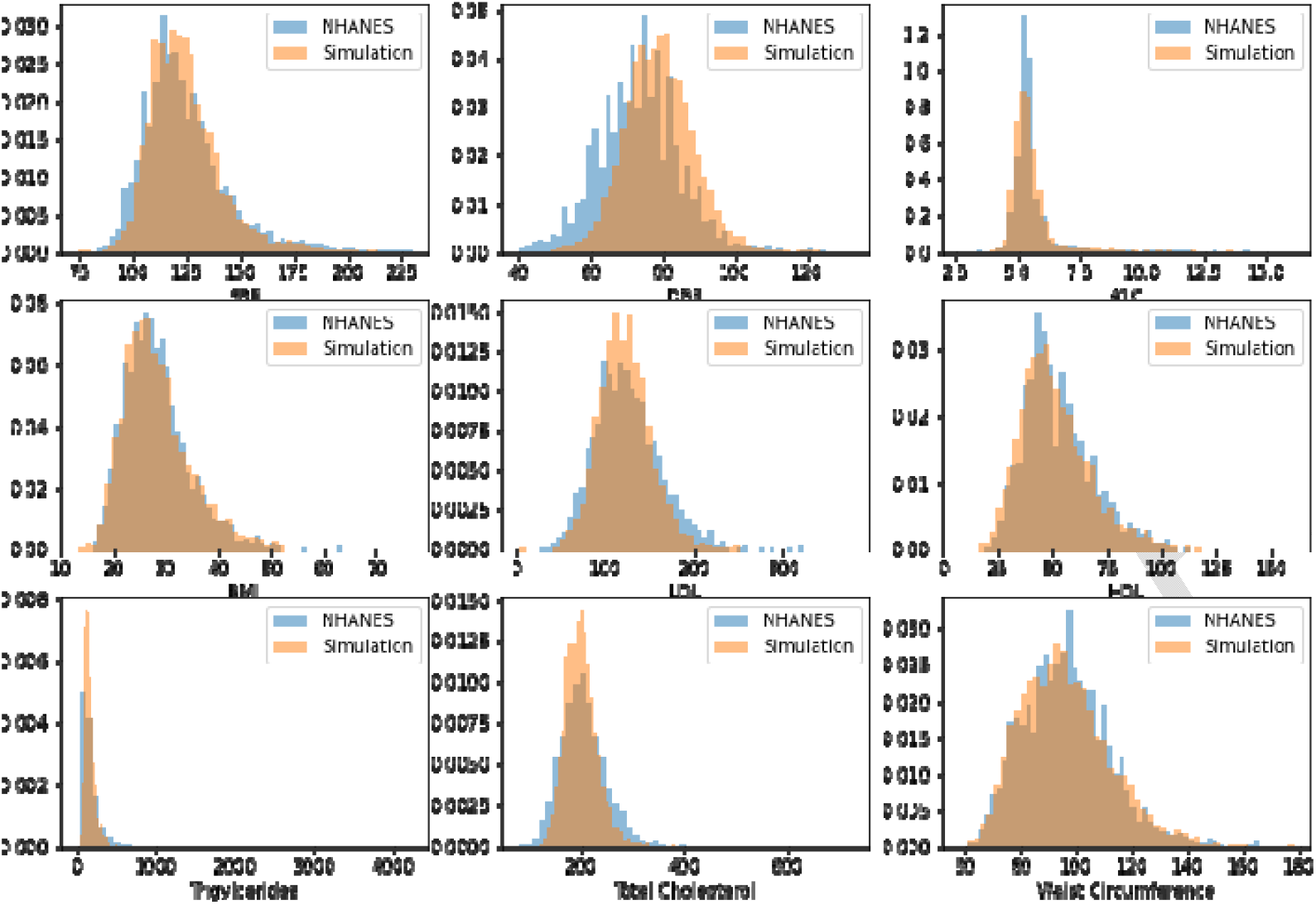
Histograms of risk factor distributions in the simulation, representing the US population, compared to a comparable NHANES sample.

### Validation of Cardiovascular Event Incidence and Mortality^51^

**Table 3** summarizes the overall incidence of stroke and MI in a simulated population of 250,000 individuals from 1999-2015. Overall age-standardized annual MI incidence was 249/100,000 population 95% CI [244 - 253] in the simulation (initialized to 1999), broadly comparable to the incidence in the Kaiser Permanente population, which ranged from 208-284 events per 100,000 from 1999-2008^52^. Overall age-standardized stroke incidence was 160 per 100,000 [157 - 164]. A wide range of stroke incidence is reported in population-based studies over this time course, ranging from 130-400/100,000^53–55^ with hospitalization rates around 200/100,000.^56^

**Table 3:**
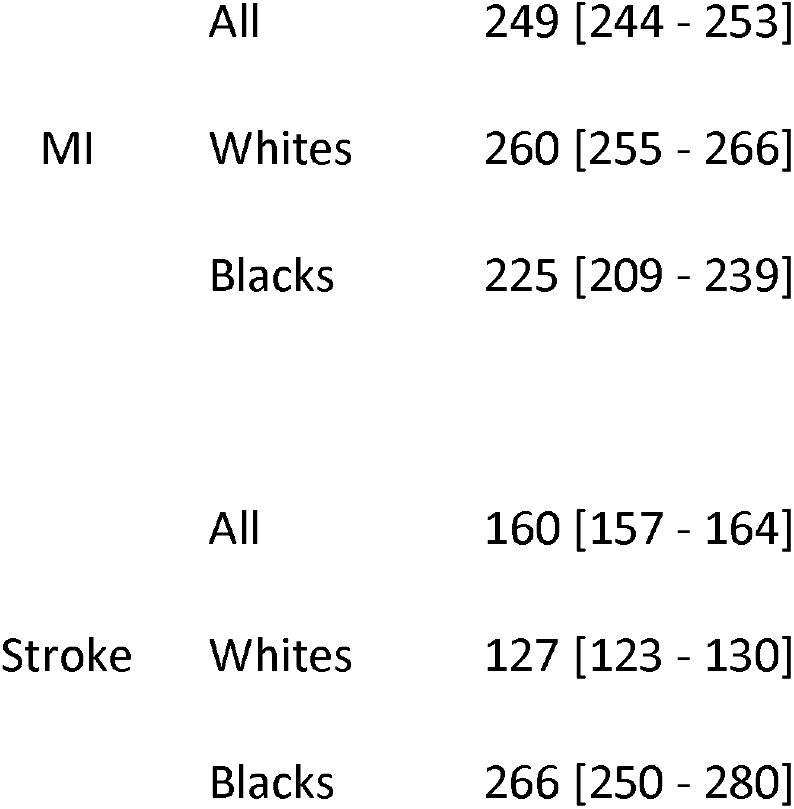
Incidence of Stroke and MI in MICROSIM (Events /100k population [95% CI] overall and by race)

Major racial disparities in stroke incidence exist, with age-standardized incidence in Black individuals generally about double the incidence in White individuals.^54,55^ In the simulation, the age-standardized incidence in Black individuals, was 266/100,000 vs. 127/100,000 in White individuals, generally reproducing reported findings in the literature. The relatively small racial differences in MI incidence reported in the literature^57^ were similarly accurately reproduced in MICROSIM’s results — incidence of 260/100,000 in White individuals vs. 225/100,000 in Black individuals.

### Validation of Treatment Effects^58^

To determine whether the simulation reproduced real-world BP treatment effects, we ran 15 simulations advancing a population of 150,000 individuals 18 years and older for 5 years under two scenarios: an “as-treated” version of the US population where BP treatment reflects current practice and a population where every individual added a single BP medication to their current regimen (mean BP lowering 5.5/3.1 mm hg). The mean RR for stroke was 0.76 [range 0.72-0.82] and the mean RR for MI was 0.85 [range 0.81-0.89]. This compares closely to our calibration standard, derived from a meta-analysis of BP-lowering treatment trials [stroke RR 0.79, MI RR 0.87].^59^

### Validation of Dementia Incidence^60^

A simulated population of 200,000 individuals 18 years and older was advanced for 20 years, and all-cause dementia incidence was tabulated. The all-cause dementia incidence within MICROSIM was then compared to the age-dementia incidence curve from the Brookmeyer et al. meta-analysis,^41^ in **Figure 3**. There was a close agreement between the MICROSIM-estimated all-cause dementia incidence and the meta-analytic all-cause dementia incidence across the age spectrum.

**Figure 3:**
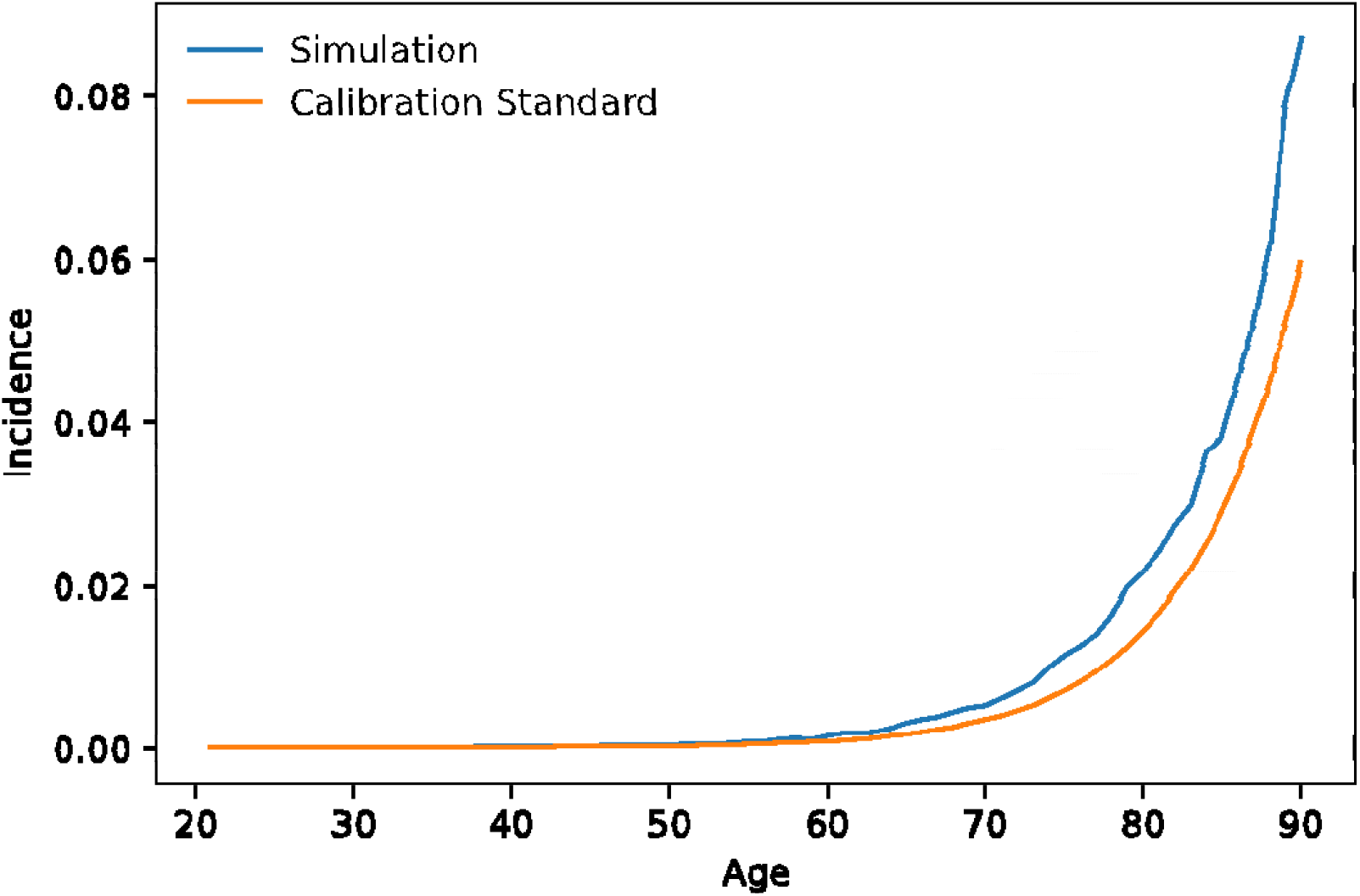
Comparison of Simulation Dementia Incidence (Incident dementia /100 population) vs. Population Standard. Legend: Comparison of raw dementia incidence by age in the simulated population vs. summarized age-specific incidence from the Brookmeyer et al, meta-analysis.

## Discussion

MICROSIM is an extensible, open-source population-based simulation model initially designed to explore cardiovascular and cognitive outcomes with varying approaches to blood pressure treatment. The core elements needed for those goals validate quite well against population-level standards. The simulation is structured to enable relatively easy modifications (e.g., changing specific risk models) and to be extended to add additional outcomes and/or structure within clinical outcomes.

In its current iteration MICROSIM has the elements in place to address a set of research questions around vascular risk factors, risk factor management, ASCVD, dementia and quality of life. It is relatively easy to slightly modify this basic framework, though, to address a variety of related research questions. For example, projecting future ASCVD and dementia under different risk assumptions and/or definitions of ASCVD and dementia or assessing the cost-effectiveness of blood pressure treatment with and without valuation of cognition. Perhaps of greater significance, the extensible framework, opens up the possibility of more comprehensively accounting for other disease states (e.g. congestive heart failure, peripheral vascular disease, chronic renal insufficiency), risk markers (e.g. social determinants of health), treatments (e.g. statins, oral hypoglycemics) and public health relevant interventions (e.g. access to nutrition, green spaces). Through such extensions, this simulation could be extended to address a vast array of research questions. In ongoing work, we are also developing frameworks to readily simulate clinical trials as well as to add disease specific phenotypic information. Specifically, we are in the process of expanding the simulation to specifically assess post-stroke vascular management by including stroke type, ischemic stroke subtypes and stroke severity.

MICROSIM’s core strengths are the application of optimal software design principles in its development, the use of the best available data to derive core simulation assumptions, and external validation of core elements. The application of programming best practices is, to our knowledge, relatively unique for an academic simulation, and we believe this will result in a sufficiently reliable codebase to generate accurate population-level inferences.

MICROSIM’s core limitations are that, as with any simulation, the results are only as strong as the core model assumptions. Given its complexity, MICROSIM relies on many assumptions that are difficult to directly evaluate given the current state of evidence (e.g., the effect of blood pressure treatment on cognition). As stronger evidence emerges, our intent is to continue to incorporate the best available evidence into MICROSIM. Similarly, the model structure assumes relatively simple relationships when reality may be more complex (e.g., the potential J-shaped curve relating DBP to mortality). Thus, optimal application of MICROSIM to specific research questions will require consideration of how those assumptions may influence results, and when uncertain model assumptions can plausibly influence conclusions, robust sensitivity analyses be conducted by altering those assumptions and reassessing results. Additionally, while race was included in the regression models underling MICROSIM, the data used to derive these regression models was often insufficient to address effects of Hispanic ethnicity (e.g. on cognition) and thus, while Hispanics are included in the model, it is not clear that MICROSIM would reproduce society level trends.

## Data Availability

All data produced in the present work are contained in the manuscript

## Cohort Funding/Support

The Atherosclerosis Risk in Communities (ARIC) study has been funded in whole or in part with Federal funds from the National Heart, Lung, and Blood Institute, National Institutes of Health, Department of Health and Human Services, under Contract nos. (HHSN268201700001I, HHSN268201700002I, HHSN268201700003I, HHSN268201700005I, HHSN268201700004I). Neurocognitive data are collected by U01 2U01HL096812, 2U01HL096814, 2U01HL096899, 2U01HL096902, 2U01HL096917, and R01 AG040282 from the NIH (NHLBI, NINDS, NIA, and NIDCD). The authors thank the staff and participants of the ARIC study for their important contributions.

The Coronary Artery Risk Development in Young Adults Study (CARDIA) is conducted and supported by the National Heart, Lung, and Blood Institute (NHLBI) in collaboration with the University of Alabama at Birmingham (HHSN268201800005I & HHSN268201800007I), Northwestern University (HHSN268201800003I), University of Minnesota (HHSN268201800006I), and Kaiser Foundation Research Institute (HHSN268201800004I). A neurocognitive ancillary study is funded by NIH (NIA, R01 AG063887).

This manuscript has been reviewed by CARDIA for the scientific content.

The Cardiovascular Health Study (CHS) was supported by contracts HHSN268201200036C, HHSN268200800007C, HHSN268201800001C, N01HC55222, 379 N01HC85079, N01HC85080, N01HC85081, N01HC85082, N01HC85083, N01HC85086, 75N92021D00006, and grants

U01HL080295 and U01HL130114 from the National Heart, Lung, and Blood Institute (NHLBI), with additional contribution from the National Institute of Neurological Disorders and Stroke (NINDS). Additional support was provided by R01AG023629, R01AG15928, and R01AG20098 from the National Institute on Aging (NIA). A full list of principal CHS investigators and institutions can be found at CHS-NHLBI.org. The content is solely the responsibility of the authors and does not necessarily represent the official views of the National Institutes of Health.

The Framingham Heart Study is a project of the National Heart Lung and Blood Institute of the National Institutes of Health and Boston University School of Medicine. This project has been funded in whole or in part with Federal funds from the National Heart, Lung, and Blood Institute, National Institutes of Health, Department of Health and Human Services, under contract No. HHSN268201500001I.

The Northern Manhattan Stroke study has been funded at least in part with federal funds from the National Institutes of Health, National Institute of Neurological Disorders and Stroke by R01 NS29993.

The Multi-Ethnic Study of Atherosclerosis (MESA) was supported by contracts 75N92020D00001, HHSN268201500003I, N01-HC-95159, 75N92020D00005, N01-HC-95160, 75N92020D00002, N01-HC-95161, 75N92020D00003, N01-HC-95162, 75N92020D00006, N01-HC-95163, 75N92020D00004, N01-HC-95164, 75N92020D00007, N01-HC-95165, N01-HC-95166, N01-HC-95167, N01-HC-95168, and N01-HC-95169 from the National Heart, Lung, and Blood Institute, and by grants UL1-TR-000040, UL1-TR-001079, and UL1-TR-001420 from the National Center for Advancing Translational Sciences (NCATS). Cognitive testing at Exam 6 in MESA has been funded by grants R01HL127659 from the National Heart, Lung, and Blood Institute (NHLBI) and R01AG054069 from the National Institutes of Health. The authors thank the other investigators, the staff, and the participants of the MESA study for their valuable contributions. A full list of participating MESA investigators and institutions can be found at http://www.mesa-nhlbi.org.

## References

1. Heller, D. J. et al. Evaluating the Impact and Cost-Effectiveness of Statin Use Guidelines for Primary Prevention of Coronary Heart Disease and Stroke. Circulation 136, 1087–1098 (2017).

2. Sussman, J., Vijan, S. & Hayward, R. Using benefit-based tailored treatment to improve the use of antihypertensive medications. Circulation 128, 2309–2317 (2013).

3. Hayward, R. A., Krumholz, H. M., Zulman, D. M., Timbie, J. W. & Vijan, S. Optimizing statin treatment for primary prevention of coronary artery disease. Annals of Internal Medicine 152, 69–77 (2010).

4. Hirsch, G., Homer, J., Trogdon, J., Wile, K. & Orenstein, D. Using Simulation to Compare 4 Categories of Intervention for Reducing Cardiovascular Disease Risks. Am J Public Health 104, 1187–1195 (2014).

5. Group, S. R. et al. A Randomized Trial of Intensive versus Standard Blood-Pressure Control. New Engl J Medicine 373, 2103–2116 (2015).

6. Group, S. M. I. for the S. R. et al. Effect of Intensive vs Standard Blood Pressure Control on Probable Dementia: A Randomized Clinical Trial. Jama 321, 553 (2019).

7. Hernán, M. A. & Robins, J. M. Using Big Data to Emulate a Target Trial When a Randomized Trial Is Not Available. Am J Epidemiol 183, 758–764 (2016).

8. Lipow, M. Number of Faults per Line of Code. IEEE Transactions on Software Engineering SE-8, 437–439 (1982).

9. k, B. et al. Manifesto for Agile Software Development. https://agilemanifesto.org.

10. Burke, J. F. & Copeland, L. Michigan Chronic Disease Simulation Model (MICROSIM).

11. Burke, J. F. & Copeland, L. Michigan Chronic Disease Simulation Model — Derivation and Validation Notebooks. (2021).

12. Black — The Uncompromising Code Formatter. https://black.readthedocs.io/en/stable/.

13. Beck, K. Test Driven Development: By Example. (Addison-Wesley Professional, 2011).

14. Travis CI: Test and Deploy with Confidence. https://travis-ci.org.

15. The Atherosclerosis Risk in Communities (ARIC) Study: design and objectives. The ARIC investigators. American Journal of Epidemiology 129, 687–702 (1989).

16. Lloyd-Jones, D. M. et al. The Coronary Artery Risk Development In Young Adults (CARDIA) Study JACC Focus Seminar 8/8. J Am Coll Cardiol 78, 260–277 (2021).

17. (CHS), M. for the C. H. S. R. G. et al. The cardiovascular health study: Design and rationale. Ann Epidemiol 1, 263–276 (1991).

18. Feinleib, M., Kannel, W. B., Garrison, R. J., McNamara, P. M. & Castelli, W. P. The framingham offspring study. Design and preliminary data. Prev Med 4, 518–525 (1975).

19. Collaborators, N. M. S. S. et al. Stroke incidence among white, black, and Hispanic residents of an urban community: the Northern Manhattan Stroke Study. Am J Epidemiol 147, 259–68 (1998).

20. Burke, J. F., Valdes-Sosa, P. A., Langa, K. M., Hayward, R. A. & Albin, R. L. Modeling test and treatment strategies for presymptomatic Alzheimer disease. PLoS ONE 9, e114339 (2014).

21. Burke, J. F. & Copeland, L. Build Baseline Population from NHANES. (2021).

22. (NCHS)., C. for D. C. and P. (CDC). N. C. for H. S. National Health and Nutrition Examination Survey Data. https://www.cdc.gov/nchs/nhanes/about_nhanes.htm.

23. Burke, J. F. & Copeland, L. Update Risk Factors over Time. (2021).

24. Yadlowsky, S. et al. Clinical Implications of Revised Pooled Cohort Equations for Estimating Atherosclerotic Cardiovascular Disease Risk. Ann Intern Med 169, 20 (2018).

25. Burke, J. F. & Copeland, L. Partition Stroke/MI Risk. (2021).

26. Kissela, B. M. et al. Abstract WP162: Outcomes and Long-Term Mortality After Ischemic Stroke in the Young: A Preliminary Analysis From the Greater Cincinnati/Northern Kentucky Stroke Study. Stroke 49, (2018).

27. Howard, G., Kleindorfer, D. O., Long, D. L., Cushman, M. & Howard, V. J. Abstract 52: Contributors to the Rural Excess in Stroke Mortality: The REasons for Geographic and Racial Differences in Stroke (REGARDS) Study. Stroke 48, (2017).

28. Dhamoon, M. S. et al. Long-Term Functional Recovery After First Ischemic Stroke: The Northern Manhattan Study. Stroke; a journal of cerebral circulation 40, 2805–2811 (2009).

29. Lisabeth, L. D. et al. Neurological, Functional, and Cognitive Stroke Outcomes in Mexican Americans. Stroke 45, 1096–1101 (2018).

30. Wadhera, R. K. et al. Association Between 30-Day Episode Payments and Acute Myocardial Infarction Outcomes Among Medicare Beneficiaries. Circulation Cardiovasc Qual Outcomes 11, e004397 (2018).

31. Burke, J. F. & Copeland, L. Build Non-CV Mortality Model in NHANES. (2021).

32. Levine, D. A. et al. Association Between Blood Pressure and Later-Life Cognition Among Black and White Individuals. Jama Neurol 77, 810–819 (2020).

33. Briceño, E. M. et al. Pre-Statistical Considerations for Harmonization of Cognitive Instruments: Harmonization of ARIC, CARDIA, CHS, FHS, MESA, and NOMAS. J Alzheimer’s Dis 83, 1803–1813 (2021).

34. Briceño, E. M. et al. PRE-STATISTICAL HARMONIZATION OF COGNITIVE MEASURES ACROSS SIX POPULATION-BASED COHORTS: ARIC, CARDIA, CHS, FHS, MESA, AND NOMAS. Alzheimer’s Dementia 14, P1611–P1612 (2018).

35. Levine, D. A. et al. Sex Differences in Cognitive Decline Among US Adults. Jama Netw Open 4, e210169 (2021).

36. Gross, A. L. et al. Effects of Education and Race on Cognitive Decline: An Integrative Study of Generalizability Versus Study-Specific Results. Psychol Aging 30, 863–880 (2015).

37. Samejima, F. Estimation of latent ability using a response pattern of graded scores. Psychometrika 34, 1–97 (1969).

38. Knopman, D. S. et al. Mild cognitive impairment and dementia prevalence: The Atherosclerosis Risk in Communities Neurocognitive Study. Alzheimer’s Dementia Diagnosis Assess Dis Monit 2, 1–11 (2016).

39. Lopez, O. L. et al. Evaluation of Dementia in the Cardiovascular Health Cognition Study. Neuroepidemiology 22, 1–12 (2003).

40. Satizabal, C. L. et al. Incidence of Dementia over Three Decades in the Framingham Heart Study. New Engl J Medicine 374, 523–532 (2016).

41. Brookmeyer, R., Gray, S. & Kawas, C. Projections of Alzheimer’s disease in the United States and the public health impact of delaying disease onset. American Journal of Public Health 88, 1337–1342 (1998).

42. Brookmeyer, R., Johnson, E., Ziegler-Graham, K. & Arrighi, H. M. Forecasting the global burden of Alzheimer’s disease. Alzheimer’s Dementia 3, 186–191 (2007).

43. Fuchs, F. D. & Whelton, P. K. High Blood Pressure and Cardiovascular Disease. Hypertension 75, 285–292 (2020).

44. Collaboration, T. B. P. L. T. T. Blood pressure-lowering treatment based on cardiovascular risk: a meta-analysis of individual patient data. The Lancet 384, 591–598 (2014).

45. Netuveli, G., Wiggins, R. D., Hildon, Z., Montgomery, S. M. & Blane, D. Quality of life at older ages: evidence from the English longitudinal study of aging (wave 1). J Epidemiol Commun H 60, 357 (2006).

46. Jönsson, L. et al. Patient-and Proxy-Reported Utility in Alzheimer Disease Using the EuroQoL. Alz Dis Assoc Dis 20, 49–55 (2006).

47. Burke, J. F. & Copeland, L. Describe and Compare Simulation Population to Published NHANES Standards. (2021).

48. Saydah, S. et al. Trends in cardiovascular disease risk factors by obesity level in adults in the United States, NHANES 1999-2010. Obesity 22, 1888–1895 (2014).

49. Muntner, P. et al. Trends in Blood Pressure Control Among US Adults With Hypertension, 1999-2000 to 2017-2018. Jama 324, 1190–1200 (2020).

50. Burke, J. F. & Copeland, L. Compare MICROSIM Vascular Risk Factors over time to NHANES Pseudo-cohort. (2021).

51. Burke, J. F. & Copeland, L. Compare MICROSIM CV Event Incidence and Population Mortality to Reported Standards. (2021).

52. Yeh, R. W. et al. Population Trends in the Incidence and Outcomes of Acute Myocardial Infarction. New Engl J Medicine 362, 2155–2165 (2010).

53. Morgenstern, L. B. et al. Persistent ischemic stroke disparities despite declining incidence in Mexican Americans. Ann Neurol 74, 778–785 (2013).

54. Kleindorfer, D. O. et al. Stroke Incidence Is Decreasing in Whites But Not in Blacks: A Population-Based Estimate of Temporal Trends in Stroke Incidence From the Greater Cincinnati/Northern Kentucky Stroke Study * Supplemental Material. Stroke; a journal of cerebral circulation 41, 1326–1331 (2010).

55. Howard, V. J. et al. Disparities in stroke incidence contributing to disparities in stroke mortality. Annals of Neurology 69, 619–627 (2011).

56. Ramirez, L. et al. Trends in Acute Ischemic Stroke Hospitalizations in the United States. J Am Heart Assoc 5, (2016).

57. Chi, G. C. et al. Trends in Acute Myocardial Infarction by Race and Ethnicity. J Am Heart Assoc 9, e013542 (2020).

58. Burke, J. F. & Copeland, L. Compare MICROSIM Treatment Effects to RCT Standards. (2021).

59. Turnbull, F. & Collaboration, B. P. L. T. T. Effects of different blood-pressure-lowering regimens on major cardiovascular events: results of prospectively-designed overviews of randomised trials. Lancet 362, 1527–1535 (2003).

60. Burke, J. F. & Copeland, L. Compare MICROSIM Dementia to Population-Level Dementia Estimates. (2021).

